## Supplementary for "Prioritization of new candidate genes for rare genetic diseases by a disease-aware evaluation of heterogeneous molecular networks"

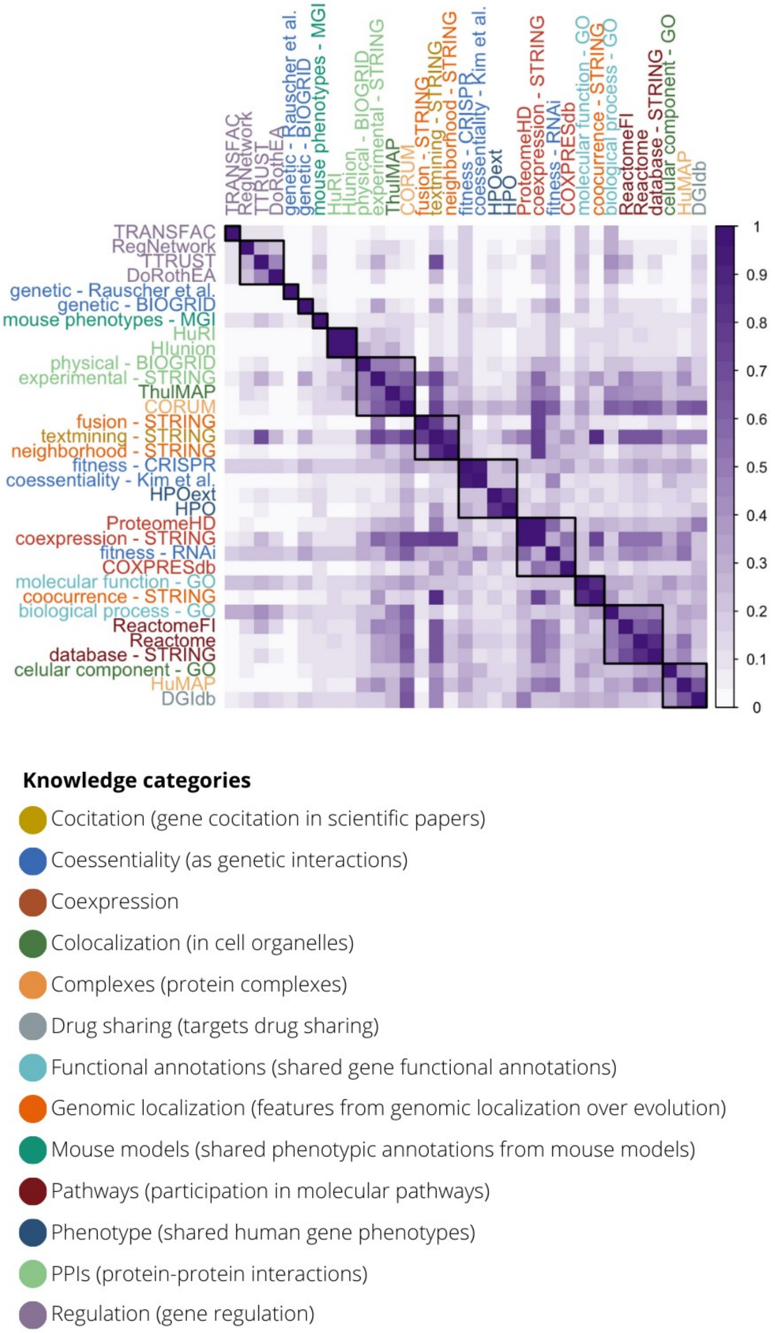

**Figure S1. Pairwise network similarity.** Edge-wise overlap coefficient is represented for each pair of networks considered in this study. Labels are colored according to the Knowledge Category (KC) in which they are classified. Hierarchical clustering of networks highlights their association by KC and reveals some networks with a global low overlap coefficient as genetic interactions from BIOGRID and, oppositely, shows networks with general high overlap such as textmining from STRING.

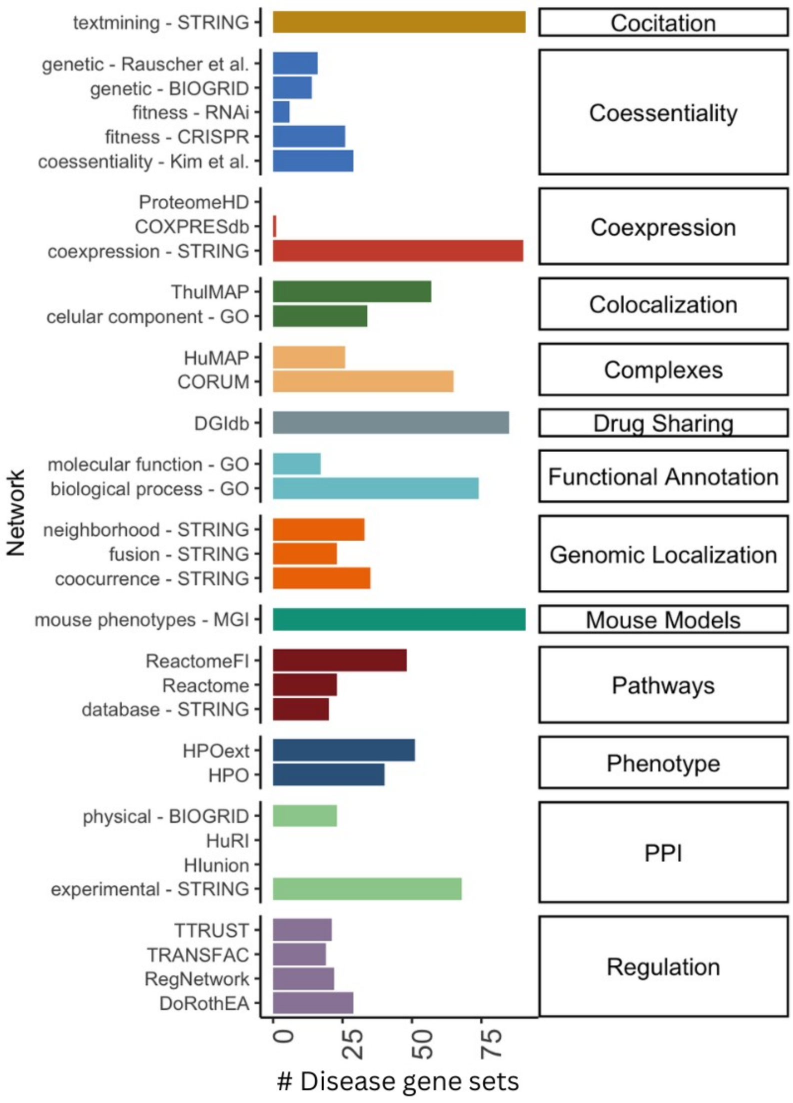

**Figure S2: Representative KC network across diseases.** For every tested disease gene set (N=91) and knowledge category (KC, N=13), we selected a single individual network based on their ability to prioritized disease genes by using their area under the precision-recall gain (AUPRG) as measure of non-random behaviour. A total of 91 disease genesets were evaluated.

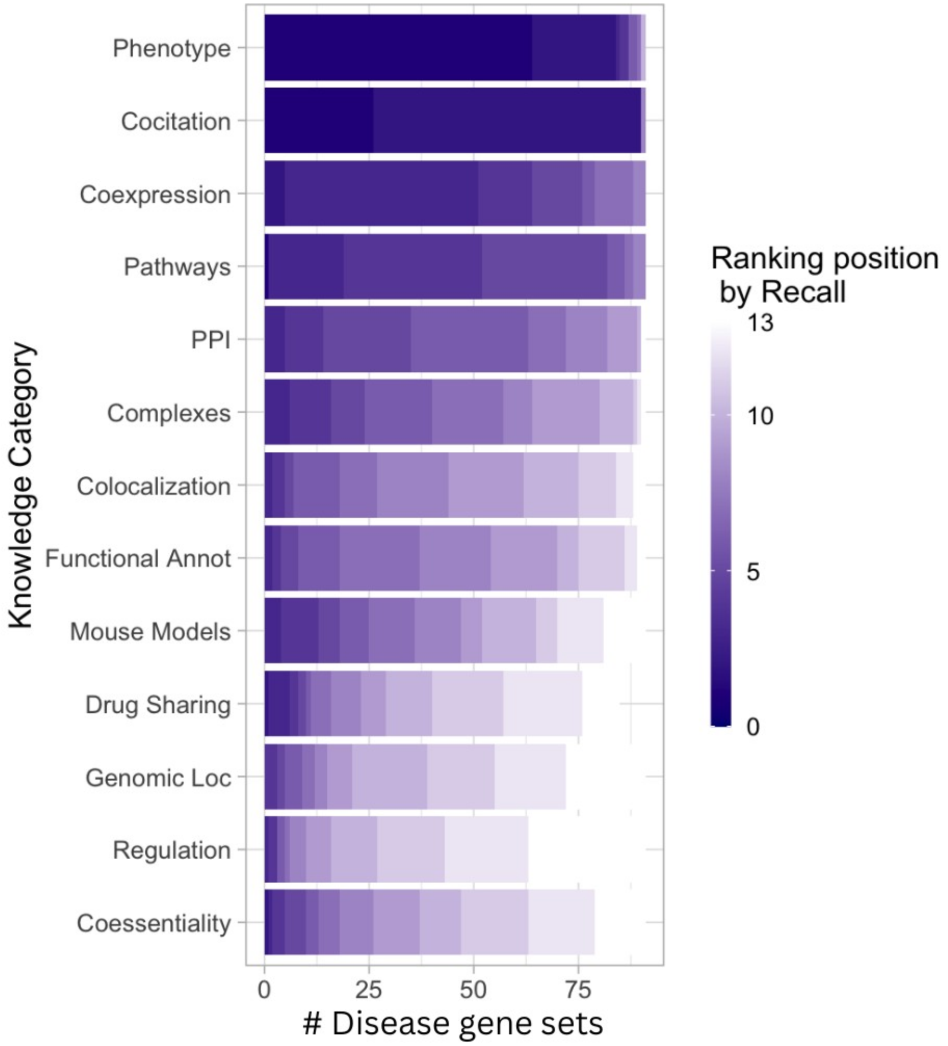

**Figure S3: Ranking of KC networks by efficiency in their recovery of gene-disease associations.** Recall at n (n = gene set size) was used as efficiency metric.

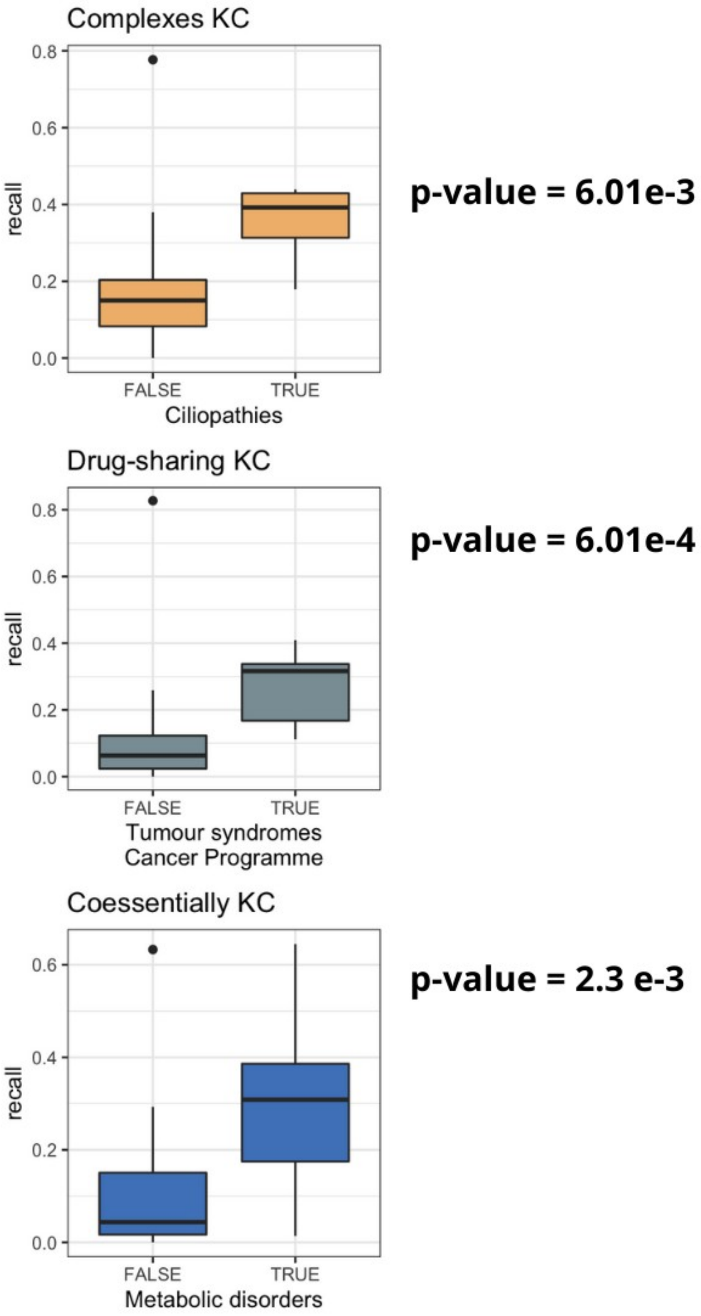

**Figure S4: High efficiency of particular KCs in specific disease families.** Recall at n-top (n = gene set size) for specific KCs are represented as a distribution classified by: TRUE (diseases of that family) and FALCE (other diseases). Increased efficiency for the disease family under study was tested using the Wilcoxon signed-rank test.

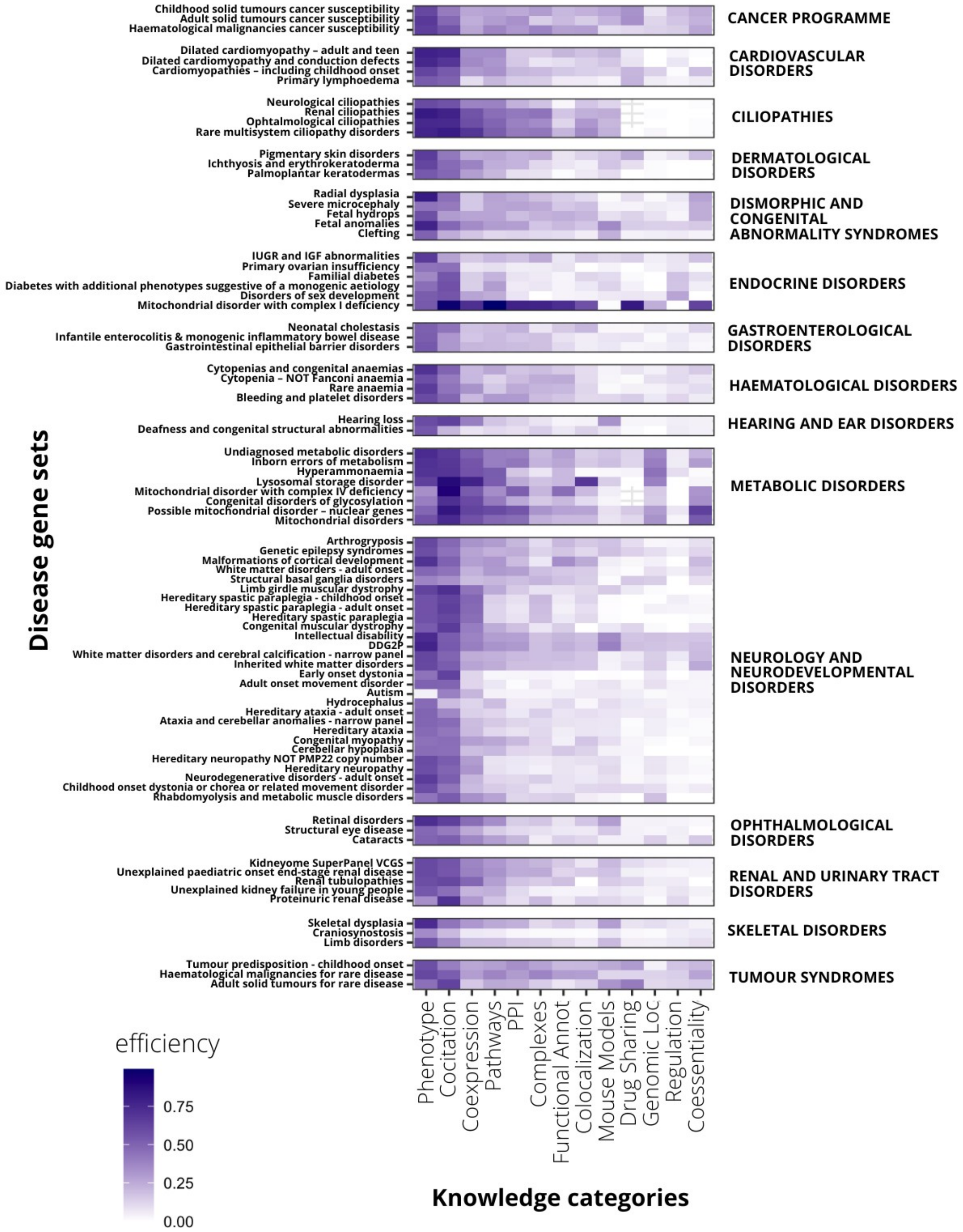

**Figure S5. Efficiency in gene recovery for every Knowledge Category (KC) in 91 diseases.** Efficiency is measured as recall at n-top (n=number of genes associated to the disease). Diseases are classified in disease families.

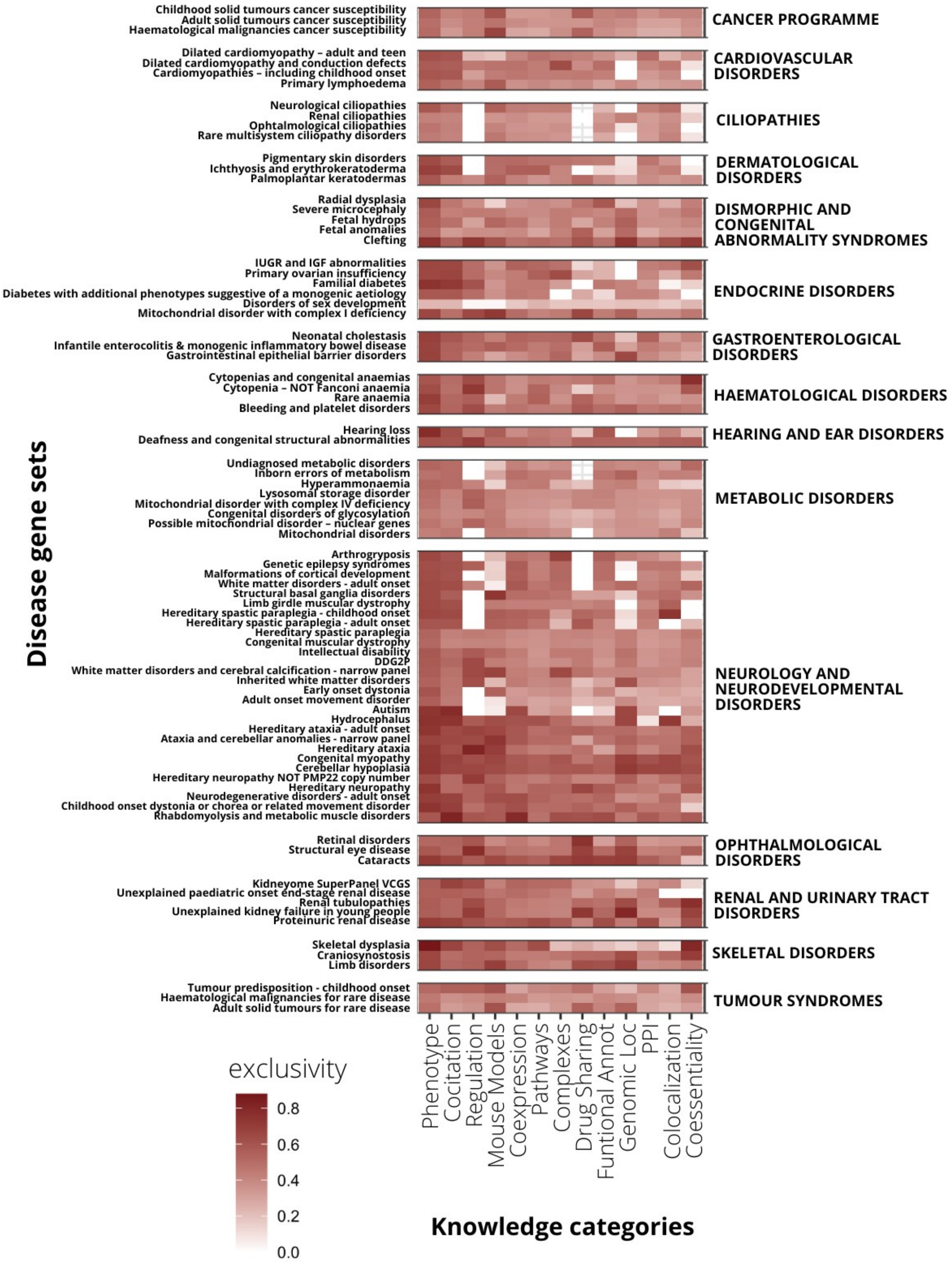

**Figure S6. Exclusivity in gene recovery for every Knowledge Category (KC) in 91 diseases.** Exclusivity is measured as the average of gene specificity at n-top (n=number of genes associated to the disease). Diseases are classified in disease families.

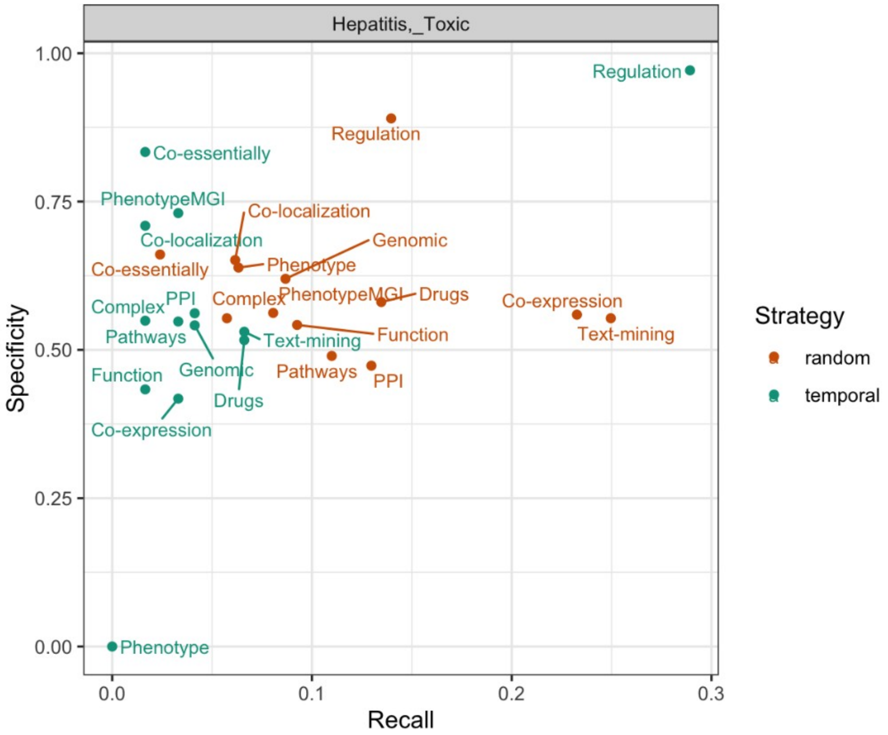

**Figure S7. KC performance across alternative network evaluation modes.** Scatterplot representing KC specificity and recall relationship for Toxic Hepatitis DisGeNET gene set using both random and time-printed network evaluation.

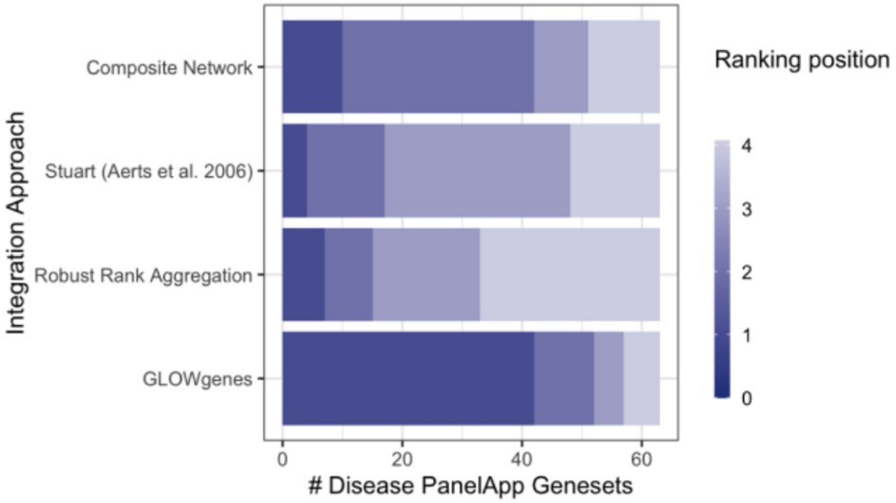

**Figure S8:** Performance ranking of alternative KC integration approaches across PanelApp disease gene sets at top-n recall (n = size of RG validation set).

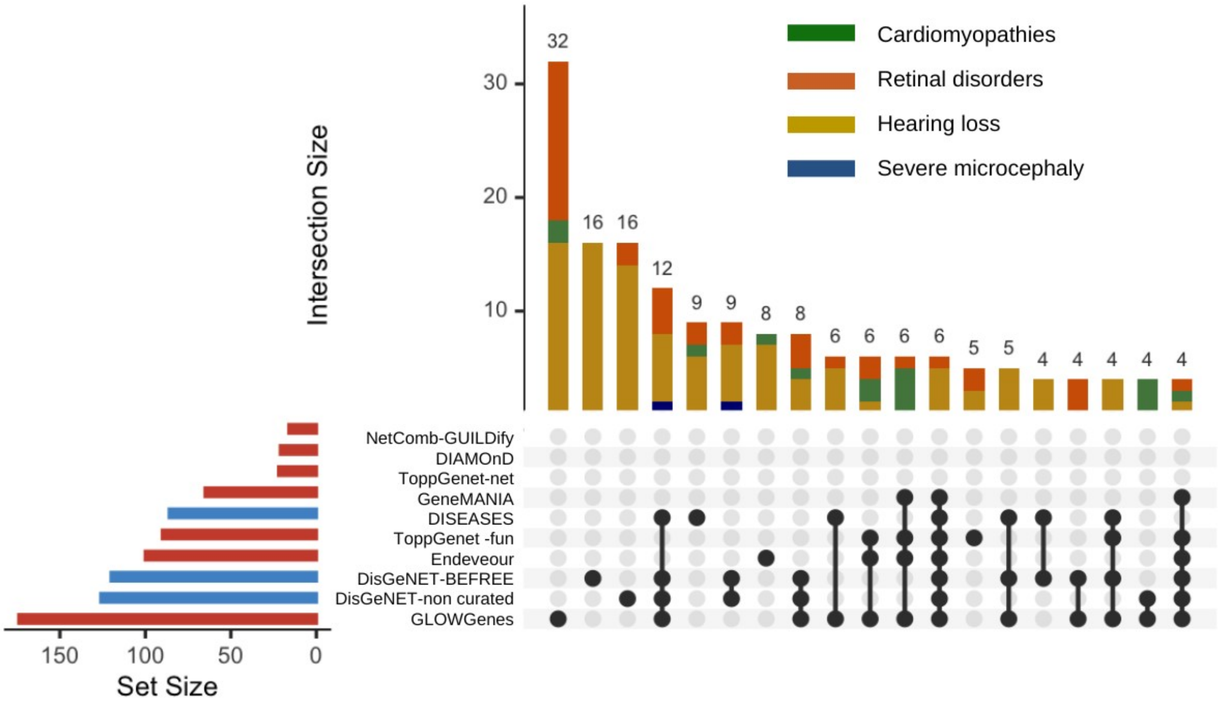

**Figure S9. Ranking of methods for prediction of gene-disease associations recovering genes from four diseases.** Number and overlap of genes captured by each of the methods evaluated at top-n (n = size of RG validation set). Diseases included are: cardiomyopathies including childhood onset, hearing loss, retinal disorders and severe microcephaly.

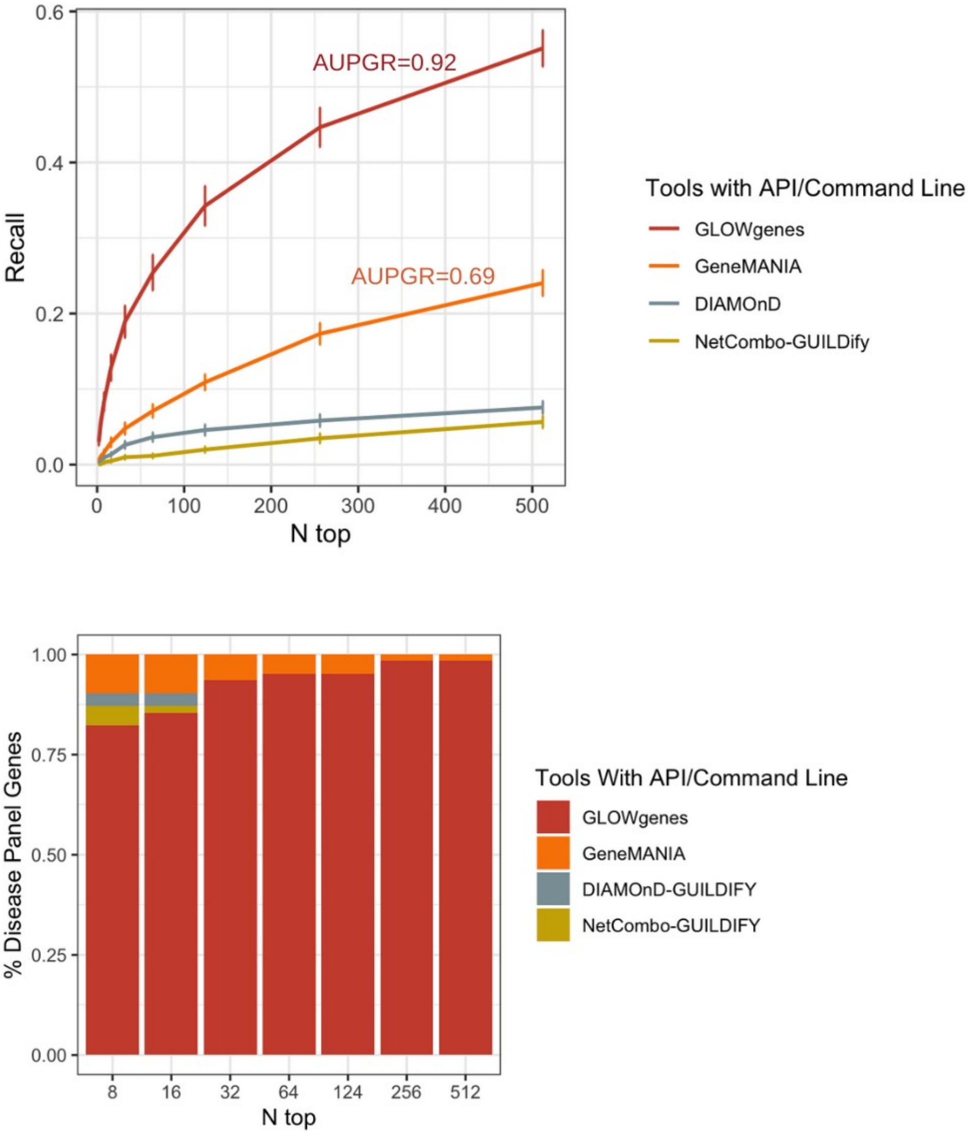

**Figure S10.** Performance ranking of gene-disease association prediction methods providing programmatic access across 70 PanelApp disease gene sets. Recall at top-n (n = size of RG validation set) was selected as threshold.

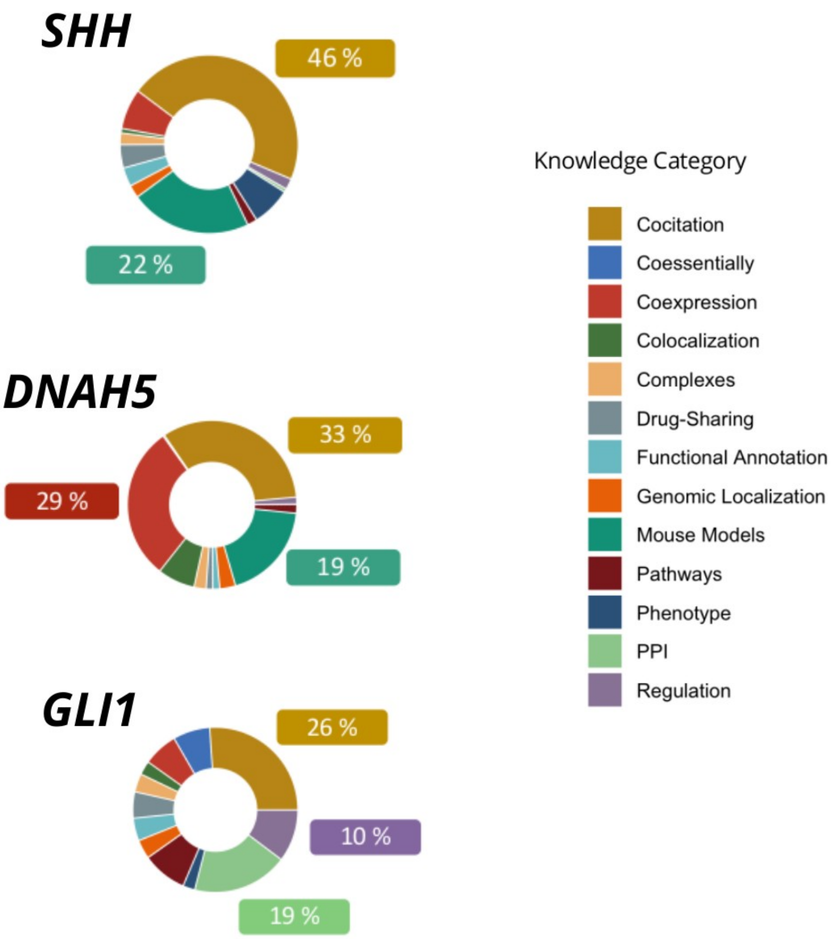

**Figure S11.** KC contribution in the prediction of three genes (*SHH*, *DNAH5* and *GLI1*) in their association of syndromic inherited retinal dystrophies. Pathogenic variants were found in this three genes in three patients.

**Table S1.** List of sources with functional information used in this study.

| Network | Knowledge Category | Edges weights | Data Source | Download Source | Reference |
|---|---|---|---|---|---|
| Textmining - STRING | COCITATION | Textmining score | STRING v11 | https://string-db.org/cgi/download.pl | Szklarczyk et al,. 2019 |
| COXPRESdb | COEXPRESSION | Inverse of the Mutual Rank value | COXPRESdb v7 - Hsa-u.c2-0 | https://coxpresdb.jp/download/Hsa-u.c2-0/ | Obayashi et al., 2019 |
| Coexpression - STRING | COEXPRESSION | Coexpression channel score | STRING v11 | https://string-db.org/cgi/download.pl | Szklarczyk et al., 2019 |
| Coregulation map | COEXPRESSION | | ProteomeHD - top-scoring 0.5% | https://www.proteomehd.net/download | Kustatscher et al.,2019 |
| Experimental subcellular map | COLOCALIZATION | | Subcellular Localization Experimental Map | http://science.sciencemag.org/highwire/filestream/694217/field_highwire_adjunct_files/9/aal3321_Thul_SM_table_S17.xlsx | Thul et al., 2017 |
| Cellular Component similarity - GO | COLOCALIZATION | Semantic similarity scores | Gene Ontology CC ontology | http://geneontology.org/docs/download-ontology/ | The Gene Ontology Consortium, 2019; Zhao et a.l. 2018; |
| Experimental - STRING | PPI | | STRING v11 - experimental channel | https://string-db.org/cgi/download.pl | Szklarczyk et al.,. 2019 |
| Physical PPI - BIOGRID | PPI | | BIOGRID 4.2 | https://downloads.thebiogrid.org/BioGRID | Oughtred et al., 2019 |
| HuRI physical PPI | PPI | | The Human Reference Interactome | http://www.interactome-atlas.org/ | Luck et al., 2020 |
| HI-union physical PPI | PPI | | The Human Reference Interactome | http://www.interactome-atlas.org/ | Luck et al,. 2020 |
| Reactome pathways | PATHWAYS | | Reactome | https://reactome.org/download-data | Jassal et al,. 2020 |
| Reactome FI | PATHWAYS | | Pathway-based functional interaction network - v2018 | https://reactome.org/about/news/133-reactomefiviz-app-version-7-2-0-released | Wu y Haw et al,. 2017 |
| Database - STRING | PATHWAYS | | STRING v11 - database channel | https://string-db.org/cgi/download.pl | Szklarczyk et al,. 2019 |
| Molecular function similarity | FUNCTIONAL ANNOTATION | Semantic similarity scores | Gene Ontology MF ontology | http://geneontology.org/docs/download-ontology/ | The Gene Ontology Consortium, 2019; Zhao et a.l. 2018; |
| Biological process similarity | FUNCTIONAL ANNOTATION | Semantic similarity scores | Gene Ontology BP ontology | http://geneontology.org/docs/download-ontology/ | The Gene Ontology Consortium, 2019; Zhao et a.l. 2018; |

| | | | | | |
|---|---|---|---|---|---|
| hu.MAP co-complex | COMPLEXES | | hu.MAP Experimental Map | http://hu.proteincomplexes.org/download | Drew et al. 2017 |
| CORUM co-complex | COMPLEXES | | CORUM | http://mips.helmholtz-muenchen.de/corum/download/coreComplexes.txt.zip | Giurgiu et al., 2019 |
| Genetic - Rauscher et al. | COESSENTIALITY | | Genetic Interaction Network from 85 CRISPR/Cas9 screens in human cancer cells | https://www.embopress.org/doi/full/10.15252/msb.20177656 | Rauscher et al. 2018 |
| Coessentiality - Kim et al. | COESSENTIALITY | weighted co-essentiality | 276 high-quality CRISPR knockout screens in cancer cell lines | https://www.ncbi.nlm.nih.gov/pmc/articles/PMC6464042/bin/LSA-2018-00278_TableS5.txt | Kim et al. 2019 |
| Fitness RNAi | COESSENTIALITY | gene fitness rank correlation coefficients | *501 cancer cell lines – RNAi - Achilles Project* | Reproduced following Pan et al. 2018 - https://figshare.com/articles/dataset/Pan_Meyers_et_al_Cell_Systems_2018_/6005297 | *Pan et al., 2018* |
| Fitness *CRISPR* | COESSENTIALITY | | *342 cancer cell lines – CRISPR-Cas - Achilles Project* | Reproduced following Pan et al. 2018 - https://figshare.com/articles/dataset/Pan_Meyers_et_al_Cell_Systems_2018_/6005297 | *Pan et al., 2018* |
| Genetic - BIOGRID | COESSENTIALITY | | BIOGRID 4.2 | https://downloads.thebiogrid.org/BioGRID | Oughtred et al., 2019 |
| DoRothEA regulation | REGULATION | evidence levels transformed into numeric values and normalized to 0-1 range. | DOROTHEA | saezlab.github.io/DoRothEA | García-Alonso et al., 2019 |
| TRANSFAC regulation | REGULATION | | TRANSFAC | NDEX | Wingender et al., 2008 |
| TTRUST regulation | REGULATION | | TTRUSTv2 | https://www.grnpedia.org/trrust/ | Han et al., 2018 |
| RegNetwork | REGULATION | | RegNetwork | http://www.regnetworkweb.org/ | Liu et al., 2015 |
| HPO similarity | PHENOTYPE | Z-scores in the distribution of Jaccard values | HPO similarity | https://hpo.jax.org/app/download/annotation | Köhler et al,. 2017 |
| Extended HPO similarity | PHENOTYPE | Z-scores in the distribution of Jaccard values | HPO similarity | https://hpo.jax.org/app/download/annotation | Köhler et al,. 2017 |
| Phenotype - MGI | MOUSE MODELS | Z-scores in the distribution of Jaccard values | MGIphenotype similarity | http://www.informatics.jax.org/downloads/reports/index.html | Bult et al., 2019 |
| DGIdb similarity | DRUG SHARING | | DGIdb Drug-Gene | http://www.dgidb.org/ | Cotto et al,. 2019 |

| | | | | | |
|---|---|---|---|---|---|
| | | | interactions | downloads | |
| Neighborhood - STRING | GENOMIC LOCALIZATION | | STRING v11- neighborhood channel | https://string-db.org/cgi/download.pl | Szklarczyk et al. 2019 |
| Coocurrence - STRING | GENOMIC LOCALIZATION | | STRING v11- coocurrence channel | https://string-db.org/cgi/download.pl | Szklarczyk et al. 2019 |
| Fusion - STRING | GENOMIC LOCALIZATION | | STRING v11- fusion channel | https://string-db.org/cgi/download.pl | Szklarczyk et al. 2019 |

**Table S2.** List of 91 gene sets from the PanelApp resource used to diagnose genetic diseases. Families taken from level 2 in PanelApp classification.

| PanelApp disease | Disease family |
|---|---|
| Childhood solid tumours cancer susceptibility | Cancer programme |
| Adult solid tumours cancer susceptibility | Cancer programme |
| Haematological malignancies cancer susceptibility | Cancer programme |
| Dilated cardiomyopathy – adult and teen | Cardiovascular disorders |
| Dilated cardiomyopathy and conduction defects | Cardiovascular disorders |
| Cardiomyopathies – including childhood onset | Cardiovascular disorders |
| Primary lymphoedema | Cardiovascular disorders |
| Neurological ciliopathies | Ciliopathies |
| Renal ciliopathies | Ciliopathies |
| Ophtalmological ciliopathies | Ciliopathies |
| Rare multisystem ciliopathy disorders | Ciliopathies |
| Pigmentary skin disorders | Dermatological disorders |
| Ichthyosis and erythrokeratoderma | Dermatological disorders |
| Palmoplantar keratodermas | Dermatological disorders |
| Radial dysplasia | Dysmorphic and congenital abnormality syndromes |
| Severe microcephaly | Dysmorphic and congenital abnormality syndromes |
| Fetal hydrops | Dysmorphic and congenital abnormality syndromes |
| Fetal anomalies | Dysmorphic and congenital abnormality syndromes |
| Clefting | Dysmorphic and congenital abnormality syndromes |
| IUGR and IGF abnormalities | Endocrine disorders |
| Primary ovarian insufficiency | Endocrine disorders |
| Familial diabetes | Endocrine disorders |

| | |
|---|---|
| Diabetes with additional phenotypes suggestive of a monogenic aetiology | Endocrine disorders |
| Disorders of sex development | Endocrine disorders |
| Mitochondrial disorder with complex I deficiency | Endocrine disorders |
| Neonatal cholestasis | Gastroenterological disorders |
| Infantile enterocolitis & monogenic inflammatory bowel disease | Gastroenterological disorders |
| Gastrointestinal epithelial barrier disorders | Gastroenterological disorders |
| Cytopenias and congenital anaemias | Haematological disorders |
| Cytopenia – NOT Fanconi anaemia | Haematological disorders |
| Rare anaemia | Haematological disorders |
| Bleeding and platelet disorders | Haematological disorders |
| Hearing loss | Hearing and ear disorders |
| Deafness and congenital structural abnormalities | Hearing and ear disorders |
| Undiagnosed metabolic disorders | Metabolic disorders |
| Inborn errors of metabolism | Metabolic disorders |
| Hyperammonaemia | Metabolic disorders |
| Lysosomal storage disorder | Metabolic disorders |
| Mitochondrial disorder with complex IV deficiency | Metabolic disorders |
| Congenital disorders of glycosylation | Metabolic disorders |
| Possible mitochondrial disorder – nuclear genes | Metabolic disorders |
| Mitochondrial disorders | Metabolic disorders |
| Arthrogryposis | Neurology and neurodevelopmental disorders |
| Genetic epilepsy syndromes | Neurology and neurodevelopmental disorders |
| Malformations of cortical development | Neurology and neurodevelopmental disorders |
| White matter disorders - adult onset | Neurology and neurodevelopmental disorders |
| Structural basal ganglia disorders | Neurology and neurodevelopmental disorders |
| Limb girdle muscular dystrophy | Neurology and neurodevelopmental disorders |
| Hereditary spastic paraplegia - childhood onset | Neurology and neurodevelopmental disorders |
| Hereditary spastic paraplegia - adult onset | Neurology and neurodevelopmental disorders |
| Hereditary spastic paraplegia | Neurology and neurodevelopmental disorders |
| Congenital muscular dystrophy | Neurology and neurodevelopmental disorders |
| Intellectual disability | Neurology and neurodevelopmental disorders |
| DDG2P | Neurology and neurodevelopmental disorders |
| White matter disorders and cerebral calcification - narrow panel | Neurology and neurodevelopmental disorders |
| Inherited white matter disorders | Neurology and neurodevelopmental disorders |
| Early onset dystonia | Neurology and neurodevelopmental disorders |

| | |
|---|---|
| Adult onset movement disorder | Neurology and neurodevelopmental disorders |
| Autism | Neurology and neurodevelopmental disorders |
| Hydrocephalus | Neurology and neurodevelopmental disorders |
| Hereditary ataxia - adult onset | Neurology and neurodevelopmental disorders |
| Ataxia and cerebellar anomalies - narrow panel | Neurology and neurodevelopmental disorders |
| Hereditary ataxia | Neurology and neurodevelopmental disorders |
| Congenital myopathy | Neurology and neurodevelopmental disorders |
| Cerebellar hypoplasia | Neurology and neurodevelopmental disorders |
| Hereditary neuropathy NOT PMP22 copy number | Neurology and neurodevelopmental disorders |
| Hereditary neuropathy | Neurology and neurodevelopmental disorders |
| Neurodegenerative disorders - adult onset | Neurology and neurodevelopmental disorders |
| Childhood onset dystonia or chorea or related movement disorder | Neurology and neurodevelopmental disorders |
| Rhabdomyolysis and metabolic muscle disorders | Neurology and neurodevelopmental disorders |
| Retinal disorders | Ophtalmological disorders |
| Structural eye disease | Ophtalmological disorders |
| Cataracts | Ophtalmological disorders |
| Kidneyome SuperPanel VCGS | Renal and urinary tract disorders |
| Unexplained paediatric onset end-stage renal disease | Renal and urinary tract disorders |
| Renal tubulopathies | Renal and urinary tract disorders |
| Unexplained kidney failure in young people | Renal and urinary tract disorders |
| Proteinuric renal disease | Renal and urinary tract disorders |
| Skeletal dysplasia | Skeletal disorders |
| Craniosynostosis | Skeletal disorders |
| Limb disorders | Skeletal disorders |
| Tumour predisposition - childhood onset | Tumour syndromes |
| Haematological malignancies for rare disease | Tumour syndromes |
| Adult solid tumours for rare disease | Tumour syndromes |
| Ehlers Danlos syndromes | Rheumatological disorders |
| Growth failure in early childhood | No family |
| Laterality disorders and isomerism | No family |
| Primary immunodeficiency | No family |
| Rare genetic inflammatory skin disorders | No family |

**Table S3:** List of gene prioritization tools used for comparative assessment.

| Name | Strategy | Method | Aggregation Approach | Training Set |
|---|---|---|---|---|
| **GLOWgenes** | search for genes associated with seeds | Network Based | Integration using ad-hoc network performance | Whole-genome |
| **Endeavour** | search for genes associated with seeds | Functional similarity | Integration using order statistics | Whole-genome |
| **ToppGenet - functional similarity** | search for genes associated with seeds | Functional similarity | Statistical meta-analysis (combined p-value) | Neighbours in a network |
| **ToppGenet - network based** | search for genes associated with seeds | Network Based | Not applied | Neighbours in a network |
| **NetComb - GUILDify** | search for genes associated with seeds | Network Based | Not applied | Whole-genome |
| **DIAMOnD** | search for genes associated with seeds | Network Based | Not applied | Whole-genome |
| **GeneMANIA** | search for genes associated with seeds | Network Based | Composite Functional Association Network | Whole-genome |
| **DisGeNET - non curated** | pre-defined disease | Gene-disease association | Aggregated Score by number and type of sources | Whole-genome |
| **DisGeNET - BEFREE** | pre-defined disease | Text-mining | Aggregated Score by number of publications | Whole-genome |
| **DISEASES** | pre-defined disease | Text-mining | Normalized number of abstracts | Whole-genome |

**Table S4:** Type of evidence sources used by prioritization methods selected for benchmark.

| | GLOW genes | Endeavour | ToppGenet - func. similarity | ToppGenet - network based | GUILDify | DIAMOnD | Gene MANIA | DisGeNET- non curated | DisGeNET- BEFREE | DISEASES |
|---|---|---|---|---|---|---|---|---|---|---|
| **Literature** | x | x | x | | | | | x | x | x |
| **Expression** | x | x | x | | | | x | | | |
| **Animal Models** | x | | x | | | | | x | | |
| **PPI** | x | x | x | x | x | x | x | | | |
| **Functional Annotations** | x | x | x | | | | | | | |
| **Regulation** | x | x | x | | | | | | | |
| **Genetic Associations** | x | x | | | | | x | x | | |
| **Phenotype** | x | x | x | | | | x | x | | |
| **Pathways** | x | x | x | | | | x | | | |
| **Genomic Loc.** | x | | | | | | | | | |
| **Complexes** | x | | | | | | | | | |
| **Chemical** | x | x | | | | | | | | |

| | | | | | | | |
|---|---|---|---|---|---|---|---|
| **information** | | | | | | | |
| **Localization** | x | x | | | | | x |
| **Sequence features** | | x | | | | | x |
| **Interologs** | | x | | | | | x |

**Table S5.** Gene set used as virtual panel in the diagnosis of syndromic retinal dystrophies in the Fundacion Jimenez Diaz University Hospital.

ABCA5, ACACB, ACAD9, ACBD5, ADCY3, ALMS1, ANAPC16, AP3D1, AP5M1, AP5Z1, ARHGEF16, ARHGEF17, ARHGEF38, ARL2BP, ARL3, ASIC5, ATP1B2, BICDL2, BPHL, BUG22, C12orf29, c16orf80, CALHM3, CASKIN1, CASZ1, CCDC51, CCP110, CCP110, CEP128, CEP162, CEP164, CEP250, CEP83, CFAP20, CFAP20, CIC, CLUAP1, CNGA3, CNGB3, COBL, COL6A6, COQ8B, COX16, CPNE1, CRTAC1, CWC27, CYP1A1, DACT1, DHX32, DHX34, DIDO1, DMBX1, DMBX1, DPP3, DSCAML1, EDEM3, EFEMP1, EIF4G3, EML4, EP300, EPB41L4A, ERICH6, FAM135B, FAM13A, FAM208B, FAM57B, FBN2, FDFT1, FDXR, FLVCR1, FRMD7, FRMPD2, GMIP, GNB1, GNPTAB, GPR45, GRK1, GRN, GTL3, GUCA1C, HEATR5A, HECTD3, HK1, HSPA9, IFT140, IGSF1, INPP5E, IPO11, IRX1, IRX5, IRX5 - IRX6, IRX6, ITIH2, KCNC2, KCNQ5, KIAA0907, KIF3A, KIF3B, KIFAB3, KIRREL2, LAMA1, LAMB2, LAMC3, LAMP1, LARGE1, LCA5L, Locus arRP, Locus LCA, LTBP1, MAP7D2, MAPRE2, MCM7, METTL9, MFRP, MFSD8, MIGA1, MYO10, MYOM1, NDUFA12, NDUFS3, NUTMD2, NXF1, OTOGL, OTX3, PANK2, PARD3, PARD3B, PCDH15, PCDHGC3, PCM1, PCYT1A, PEX6, PKM, PLA2G5, PLXNB2, PLXNB3, POMGNT2, POMZP3, PPP1R21, PQLC2, PRDM13, PRDM13-IRX1, PREX2, PRPF4B, PRPS1, PTPRN2, PWWP2A, RAB5B, RANBP2, RASGRF2, RCBTB1, RDH12, RECQL4, RGS22, RGS7, RIMS2, RNU4ATAC, ROM1, RP1, RTTN, RUSC1, SAMD7, SAP30, SASS6, SCLT1, SF3B2, SGSH, SH3GL2, SHROOM2, SLC26A7, SLC37A3, SLC4A7, SLC6A6, SNX29, SON, SSBP1, SUFU, SYTL4, TBC1D32, TBC1D5, TCF20, TECPR2, TJP1,TKTL2, TMED7, TMEM87B, TNXB, TRAPPC14, TRPM2, UBAP1L, USH2A, USP15, USP16, VCAN, VPS13B, VPS13D, VSX2, XPNPEP2
